# Effect of Criminalization of Drink-Driving on Road Traffic Mortality in China

**DOI:** 10.1101/2021.07.16.20242545

**Authors:** Feng Sha, Bingyu Li, Jinling Tang

**Affiliations:** Shenzhen Institutes of Advanced Technology, Chinese Academy of Sciences, Shenzhen, China; Law School, Shenzhen University, Shenzhen, China; Guangzhou Women and Children’s Medical Centre, Guangzhou, China; School of Public Health and Primary Care, The Chinese University of Hong Kong, Hong Kong, China

## Abstract

To evaluate the national effect of CDD on reducing road traffic mortality, we analyzed crude road traffic mortality rates data collected between 2006 and 2016 through China’s National Disease Surveillance System. Linear regression models were fit with the pre-CDD data (2006-2011) and used to predict mortality rates in the post-CDD years (2012-2016). It is estimated that the new law was associated with 317,197 (95% CI: 280,425∼353,968) lives saved in the entire country in the first 5-years of the new law. Similar reduction in mortality rates was observed in both urban and rural areas. The decline in non-occupants was more prominent and occurred earlier than that in occupants, among whom the road traffic mortality rate did not show a statistically significant reduction in the first 4 years of the new law. Our study shows that CDD is particularly effective in protecting non-occupants and is equally effective in both rural and urban areas in China.

## Introduction

Given the contribution of drink-driving to road traffic mortality in China, the Road Traffic Safety Law was revised in May 2011 to criminalize drink-driving (CDD)^1^. Since then driving with a blood alcohol concentration of 0.08 g/dl at a random breath test can be sentenced to 8-15 days in prison^1^. This is the first study to evaluate the national effect of CDD on reducing road traffic mortality, with particular focus on its protective effect on vulnerable road users such as pedestrians, cyclists and motorcyclists who account for the majority of traffic deaths in China^2^.

## Methods

We analyzed crude road traffic mortality rates data collected between 2006 and 2016 through China’s National Disease Surveillance System^2^. Road traffic mortality is referred to deaths caused by motor vehicle traffic accidents. Registered vehicles and population data were obtained from the National Bureau of Statistics^3^. Linear regression models were fit with the pre-CDD data (2006-2011) and used to predict mortality rates in the post-CDD years (2012-2016). The difference between predicted and actual mortality rates were used as the estimate of the effect of the new law. This difference is multiplied by China’s total population in a post-CDD year to estimate the number of lives saved in that year. Subgroup analyses by location (rural and urban) and type of road users (occupants and non-occupants) were also conducted.

## Results

The road traffic mortality rate increased linearly as the number of registered vehicles increased in the pre-CDD years. Although the number of vehicles had been steadily growing, a sudden decline in the mortality rate started in 2011∼2012, the year immediately after the new law was introduced. Compared with the expected RTM rate, the actual mortality rate was lowered relatively by 4.92% (95% CI: 1.60%∼8.24%) in the first year after the new law and by as much as 40.93% (95% CI: 38.05%∼43.82%) 5 years later (2016). It is estimated that the new law was associated with 317,197 (95% CI: 280,425∼353,968) lives saved in the entire country in the first 5-years of the new law.

Similar reduction in mortality rates was observed in both urban and rural areas. The decline in non-occupants was more prominent and occurred earlier than that in occupants, among whom the road traffic mortality rate did not show a statistically significant reduction in the first 4 years of the new law. Of the 317,197 lives saved, 84.66% were pedal-cyclists, motorcyclists and pedestrians, and only 4.39% were occupants.

## Discussion

The concurrence of the decline in road traffic mortality rate and the launch of the new law indicates that legislation is likely effective in regulating risky driving behaviors. Although the national consumption of alcohol has increased since 1990^4^, few drivers in China risk drink-driving now. This is also evidenced in the rising popularity of online designated driver service. A major limitation of this preliminary study is the before-and-after design. Potential confounding variables of this study include other road safety measures such as increased seat belt use that may also affect road traffic mortality rate. However, since the establishment of national seat belt law in 2004^5^, there has been no new major regulations or laws regarding road safety other than CDD in the study period. Therefore, the sudden decline in traffic mortality in 2011 is most likely the result of CDD. As previously found in China, the road traffic mortality rate is disproportionately higher among non-occupants than car occupants, and is also higher in rural areas than urban ones^2^. Our study shows that CDD is particularly effective in protecting non-occupants and is equally effective in both rural and urban areas in China.

## Data Availability

Data are from the below published study:
Wang L, Ning P, Yin P, Cheng P, Schwebel DC, Liu J, Wu Y, Liu Y, Qi J, Zeng X, Zhou M. Road traffic mortality in China: analysis of national surveillance data from 2006 to 2016. The Lancet Public Health. 2019 May 1;4(5):e245-55.

## Authors’ Contributions

Prof. Tang had full access to all of the data of the study and takes responsibility for the integrity of the data and the accuracy of the data analysis.

Concept and design: All authors.

Acquisition, analysis, or interpretation of data: Sha, Tang. Drafting of the manuscript: Sha, Li.

Critical revision of the manuscript for important intellectual content: All authors. Statistical analysis: Sha. Supervision: Tang.

---

**Figure.**
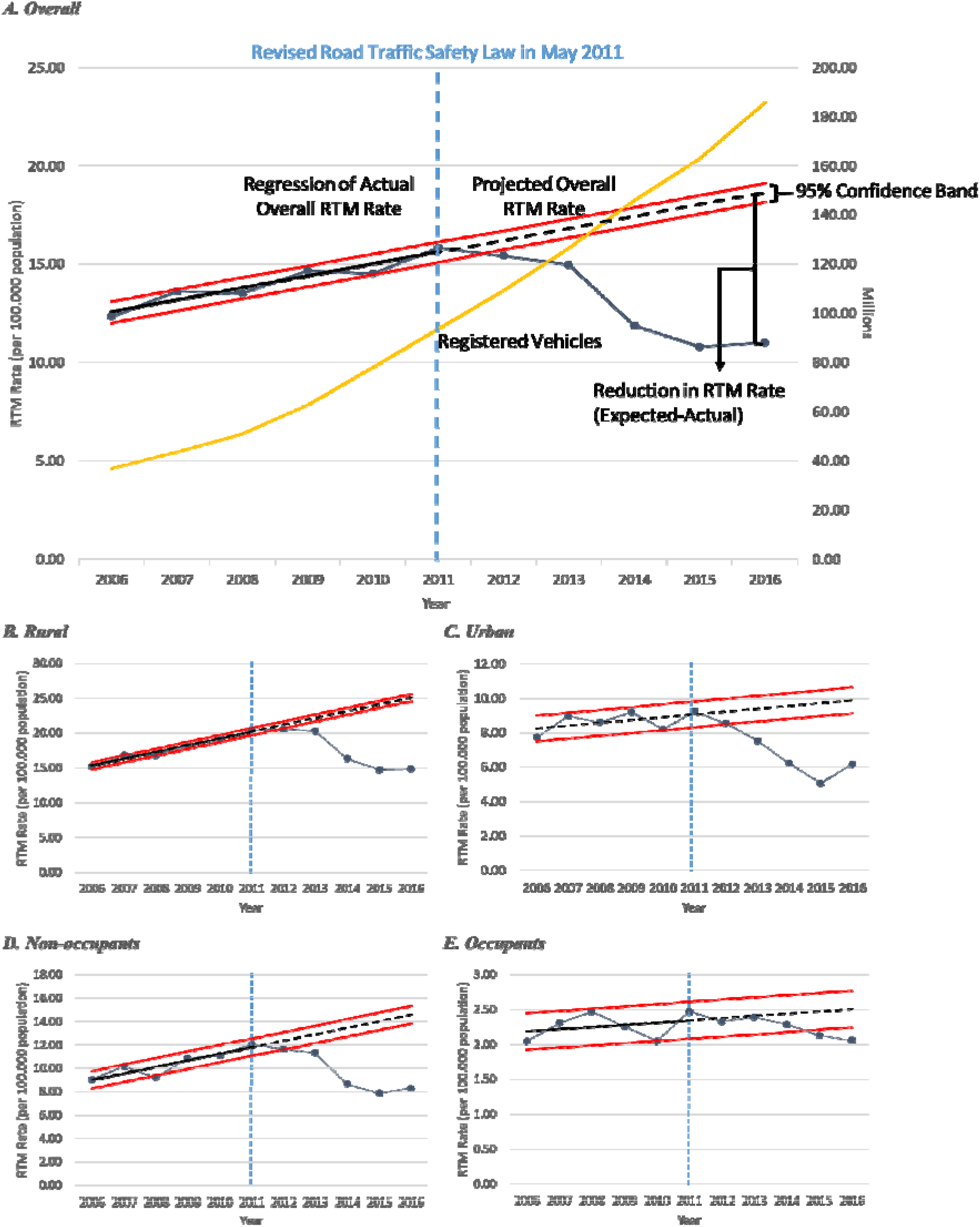
Road Traffic Mortality (RTM) Rate and Registered Vehicles in the Years before and after the New Law. Note: 1. Overall mortality rate =100,000×(the total number of traffic deaths / China’s total population in the same year). 2. Rural mortality rate = 100,000×(the number of traffic deaths / China’s total population in the rural areas in the same year); Urban mortality rate was computed in the same way. 3. Occupant mortality rate =100,000× (the number of traffic deaths in occupants / China’s total population in the same year); Non-occupant mortality rate was computed in the same way.

